# Lifetime exposure to violence and early cardiometabolic risk factors in a healthy Swedish cohort

**DOI:** 10.1101/2023.10.26.23297648

**Authors:** Rebekka Lynch, Thor Aspelund, Fang Fang, Jacob Bergstedt, Arna Hauksdóttir, Filip K. Arnberg, Þórdís Jóna Hrafnkelsdóttir, Nancy L. Pedersen, Unnur Valdimarsdóttir

## Abstract

**Introduction:** Violence exposure has been associated with cardiovascular disease. Less is known about underlying mechanisms, including early cardiometabolic risk factors, and possible sex differences of such associations.

**Methods:** We used data from the Swedish LifeGene study on 23,215 males and females, aged 18-50 years. At baseline (2009-1016) participants answered the Life Stressor Checklist-Revised alongside questions on medical diagnoses of hypertension, diabetes, dyslipidemia and smoking history. At a clinical visit, blood pressure, BMI, glycated hemoglobin (HbA1c), total cholesterol, ApoB/ApoA1 ratio, and high-sensitivity C-Reactive Protein (hs-CRP) were measured. Modified Poisson and linear regression were used to test the association between violence and cardiometabolic risk factors.

**Results:** At mean age 33±8 years, lifetime exposure to violence was reported by 23% of females and 15% of males. Those exposed to violence were more likely to smoke (PR 1.86, CI: 1.66–2.07) and report a diagnosis of hypertension (PR 1.39, CI: 1.18-1.64). While no differences were observed in measured systolic blood pressure (B -0.34, CI: -0.70, 0.02), HbA1c (B 0.06, CI: - 0.08, 0.20) or total cholesterol (B -0.01, CI: -0.04, 0.02), both males and females exposed to violence had higher BMI (B 0.51, CI: 0.39–0.63) and hs-CRP (B 0.11, CI: 0.06–0.16), after adjustment. Violence in childhood, as opposed to adulthood, and exposure to both sexual and physical violence, as opposed to either type, was more strongly associated with hs-CRP and BMI.

**Discussion:** In a young healthy Swedish sample, lifetime exposure to violence was associated with some but not all early cardiometabolic risk factors among both males and females.

## Introduction

Cardiovascular disease is one of the leading causes of death[1] and disability[1] in the European Union. Risk factors for cardiovascular disease traditionally include cardiometabolic markers, such as obesity, hypertension, diabetes, and smoking, as well as inflammation[2]. Psychological factors have been associated with cardiovascular disease, including traumatic life events such as violence[3], a potent traumatic stressor that affects almost a third of women worldwide[4].

Research on the association between violence and cardiovascular disease has focused on childhood maltreatment, including exposure to physical and sexual abuse. Studies on childhood maltreatment consistently show an association with cardiovascular disease[5] though differences by gender and sex are underreported as most studies focus on women and, in addition, use self-reported outcomes. Studies on adult exposure to violence such as intimate partner violence (which more often occurs among women[6]) are few. A recent study using UK registry data found a positive association between being exposed to domestic abuse and cardiovascular mortality among women[7]. Others have found positive self-reported associations[8, 9], though only one study examining this association included both genders and it found no association among men[10]. Conversely, men are overrepresented in military cohorts, where combat veterans have been found to have a higher odds of being diagnosed with coronary heart disease[11], compared with other deployed veterans without combat exposure.

Cardiovascular disease has a complex pathogenesis, with many risk factors and intermediaries being termed cardiometabolic factors. Obesity[12, 13], hypertension[7, 14-16], dyslipidemia[16], and diabetes[7, 17, 18], as well as inflammation[19, 20] are common cardiometabolic factors that have been studied with regard to violence exposure. Additionally, psychological factors such as depression and substance use disorder have been associated with both exposure to violence and cardiovascular disease[21, 22]. The largest cohort studying the association between violence and cardiometabolic factors is the U.S. based all-female Nurses’ Health Study II, reporting an association between childhood abuse or intimate partner violence and unhealthy eating[23], hypertension[14, 15], and diabetes[17, 18]. To our knowledge, it is the only cohort that has used validated self-reported outcomes for a range of cardiometabolic factors, as well as inflammation[19], though it only examines women, and needs replicating.

Leveraging the relatively young and healthy Swedish LifeGene cohort of both sexes, our aim is to explore the association between lifetime exposure to violence and early cardiometabolic risk factors, measured through self-reports and objective measurements among relatively young males and females. A secondary aim is to examine these associations by timing of violence, that is whether it occurred in childhood or adulthood, as well as categorizing the type of violence, that is physical or sexual, with exposure to both being used as a proxy for severity.

## Methods

### Study population

We used the Swedish LifeGene cohort, a large cohort that aims to study gene-environment interactions prospectively, with a special focus on cardiovascular disease[24]. The study recruited individuals aged 18-50 years, randomly sampled from the general population, with the option to further invite family members in the same household, both minors and adults (irrespective of age). Participants electronically signed an informed consent by logging onto a website where they thereafter responded to a comprehensive online questionnaire. This was followed by an appointment at a test center for diverse physiological measurements, as well as blood and urine collection. Data used here is from 23,215 individuals that completed the baseline assessment, collected from 2009 to 2016, and who answered at least one question of the Life Stressor Checklist-Revised, to ascertain exposure, and attended a clinical visit at the test center. The study was approved by the Regional Ethics Committee in Stockholm (Ref. no. 2018/1152-31/2).

### Exposure ascertainment

Exposure to violence was assessed during the web-based survey using the Life Stressor Checklist - Revised (LSC-R)[25]. The 30-item yes/no checklist includes a range of experiences and has seven questions specifically on violence (Supplementary Table 1). Answering at least one of these questions affirmatively classifies the person as exposed, whereas others are assumed unexposed to violence. These questions were then used to categorize the type of violence, that is exposure to only physical violence), only sexual violence, or exposure to both physical and sexual violence. Age at first exposure was then ascertained by using the first six questions on violence, with any exposure before 18 defining childhood exposure. Those only exposed as adults were categorized as exposed in adulthood. The web-based design was constructed so that if the checklist was filled out, every question had to be answered, resulting in no missing data on the exposure ascertainment.

### Outcome ascertainment

The outcomes of this study are various cardiometabolic markers, both self-reported and objectively measured, as well as mental health indicators.

Participants self-reported diagnoses of hypertension, diabetes type 2, hyperlipidemia, and current smoking. Individuals who endorsed drinking at least monthly were asked to answer the Fast Alcohol Screening Test (FAST)[26], a shortened version of the Alcohol Use Disorders Identification Test (AUDIT) to screen for possible alcohol use disorder. It contains four items and is considered positive for possible alcohol use disorder if three or more items are endorsed. All participants were asked to answer the self-reported version of WHO’s Composite International Diagnostic Interview’s module on depression[27], with positive depressive symptoms if five or more questions were answered affirmatively.

At the clinical visit to the test center, the segmental body composition analyzer Tanita BC-418 MA was used to assess body mass index (BMI). Sitting systolic and diastolic blood pressure readings (mmHg) were obtained manually using the right arm at rest, with measurements >140/90 being retaken. Participants gave a blood sample (EDTA whole blood) that was used to measure total cholesterol (mmol/L), glycated hemoglobin (HbA1c, mmol/mol), high-sensitivity C-reactive protein (hs-CRP, mg/L) and Apolipoprotein A1 and B (g/L), in two different laboratories in Stockholm, Sweden. This information was then used to calculate the ratio between Apolipoprotein B and Apolipoprotein A1 (ApoB/ApoA1 ratio). Hs-CRP values over 10 mg/L were excluded (N = 259, 25% with violence exposure), as such high levels may be related to acute infection or recent physical trauma. 5,417 individuals had undetectable levels of hs-CRP in the assay. We imputed those individuals with a value 50% lower than the lowest detection level, that is 0.1mg/L.

### Covariates

Age was determined at the test center visit. Educational level was categorized as primary, secondary, and university, by highest level attended. Occupation was divided into three categories: employed, student, and other (unemployed, disability, retired, homemaker, sick leave). Civil status categories were created by merging marriage and cohabitation as one category, single and widowed as a second category, and in a relationship as a third category.

### Statistical analysis

Background characteristics are described using frequency and means with standard deviation by exposure to violence overall, as well as with exposure status by sex. Prevalence ratios (PR) with 95% confidence intervals (CI) for dichotomized outcomes were calculated using a modified Poisson regression with a sandwich variance estimator[28] with adjustment for age (continuous) and sex (whole sample) in a simpler model, or with adjustment for age, occupation, marital status, and sex in an extended model. Linear regression models were used for continuous data, with the same adjustments in two models. We estimated sex-specific associations in both the simple and extended model by stratifying on sex. The exposure was analyzed as a binary yes/no and then as a categorical variable with levels: unexposed, first exposed in childhood, and first exposed in adulthood. We also analyzed by category of violence exposure (physical violence only, sexual violence only, and both physical and sexual violence), using estimated marginal means to compare these categories when confidence intervals suggested a statistical difference[29]. All calculations were performed in R v4.0.3.

## Results

Among 23,215 participants, 4,688 (20.2%) had experienced violence during their lifetime, with a higher prevalence among females (23.3% vs. 15.5% among males). Females were more likely to have experienced sexual violence (15.8% vs. 2.5%) and slightly less likely to report history of physical violence (11.6% vs. 14.0%) compared with males (Supplementary Table 1). Table 1 describes the background characteristics of the participants. Mean age was a year higher among individuals exposed to violence compared with unexposed (33.9 ± 8.1 vs 32.9 ± 8.1). The majority of participants had attended university, were employed and married/cohabitating. Overall, 78.2% of participants had a blood sample collected, with a slightly higher proportion of exposed individuals with a blood sample (79.4% vs. 78.0%). Individuals who gave a blood sample were more likely to have a university education but other characteristics were similar (Supplementary Table 2).

**Table 1.** Background characteristics of the participants by sex and violence exposure in the Swedish Lifegene Cohort.

|  | All |  | Females |  | Males |  |
| --- | --- | --- | --- | --- | --- | --- |
|  | Unexposed<br>N = 18527 | Violence<br>N = 4688 | Unexposed<br>N = 10757 | Violence<br>N = 3268 | Unexposed<br>N = 7770 | Violence<br>N = 1420 |
| **Age, mean (SD)<sup>1</sup>** | 32.9 (8.1) | 33.9 (8.1) | 32.5 (8.2) | 33.7 (8.2) | 33.5 (7.9) | 34.3 (7.6) |
| Information missing | 0 (0.0) | 0 (0.0) | 0 (0.0) | 0 (0.0) | 0 (0.0) | 0 (0.0) |
| **Education level (%)** |  |  |  |  |  |  |
| Primary | 188 (1.6) | 101 (2.2) | 79 (0.7) | 66 (2.0) | 109 (1.4) | 35 (2.5) |
| Secondary | 5016 (27.1) | 1556 (33.2) | 2734 (25.4) | 1051 (32.2) | 2282 (29.4) | 505 (35.6) |
| University | 13081 (70.6) | 2995 (63.9) | 7816 (72.7) | 2126 (65.1) | 5265 (67.8) | 869 (61.2) |
| Information missing | 242 (1.3) | 36 (0.8) | 128 (1.2) | 25 (0.8) | 114 (1.5) | 11 (0.8) |
| **Occupation (%)** |  |  |  |  |  |  |
| Employed | 12943 (69.9) | 3192 (68.1) | 7286 (67.7) | 2158 (66.0) | 5657 (72.8) | 1034 (72.8) |
| Student | 2821 (15.2) | 688 (14.7) | 1784 (16.6) | 517 (15.8) | 1037 (13.3) | 171 (12.0) |
| Other<sup>2</sup> | 738 (4.0) | 345 (7.4) | 456 (4.2) | 259 (7.9) | 282 (3.6) | 86 (6.1) |
| Information missing | 2025 (10.9) | 463 (9.9) | 1231 (11.4) | 334 (10.2) | 794 (10.2) | 129 (9.1) |
| **Civil status (%)** |  |  |  |  |  |  |
| Marriage/Cohabitation | 11210 (60.5) | 2712 (57.8) | 6489 (60.3) | 1895 (58.0) | 4721 (60.8) | 817 (57.5) |
| Relationship | 1766 (9.5) | 520 (11.1) | 1066 (9.9) | 375 (11.5) | 700 (9.0) | 145 (10.2) |
| Single/Widowed | 5178 (27.9) | 1397 (29.8) | 2996 (27.9) | 956 (29.3) | 2182 (28.1) | 441 (31.1) |
| Information missing | 373 (2.0) | 59 (1.3) | 206 (1.9) | 42 (1.3) | 167 (2.1) | 17 (1.2) |
| **Blood draw performed (%)** | 14442 (78.0) | 3721 (79.4) | 8371 (77.8) | 2571 (78.7) | 6071 (78.1) | 1150 (81.0) |

<sup>1</sup> Standard deviation
<sup>2</sup> Unemployed, disability, retired, homemaker or sick leave

Figure 1 and Supplementary Table 3 show the prevalence ratios of cardiometabolic diagnoses and mental health indicators by exposure to violence. Individuals exposed to violence were, compared with unexposed individuals, more likely to smoke (PR 1.86; CI: 1.66, 2.07) and report a diagnosis of hypertension (PR 1.39; CI: 1.18, 1.64). There were no sex differences in these associations after covariate adjustment. No statistically significant associations were found in the prevalence of diabetes type 2 (0.2% vs 0.1%) or hyperlipidemia (1.9% vs 1.6%) diagnoses. There was a higher prevalence of obesity among individuals exposed to violence (PR 1.46; CI: 1.28, 1.66, Supplementary Table 3), after adjustment, with little sex differences. Those exposed to violence were more likely to have possible alcohol use disorder (PR 1.45; CI:1.30, 1.61) and experience depressive symptoms (PR 1.99; CI: 1.87, 2.11) after adjustment. Similarly, we found no evidence of sex differences in these associations.

**Figure 1.**
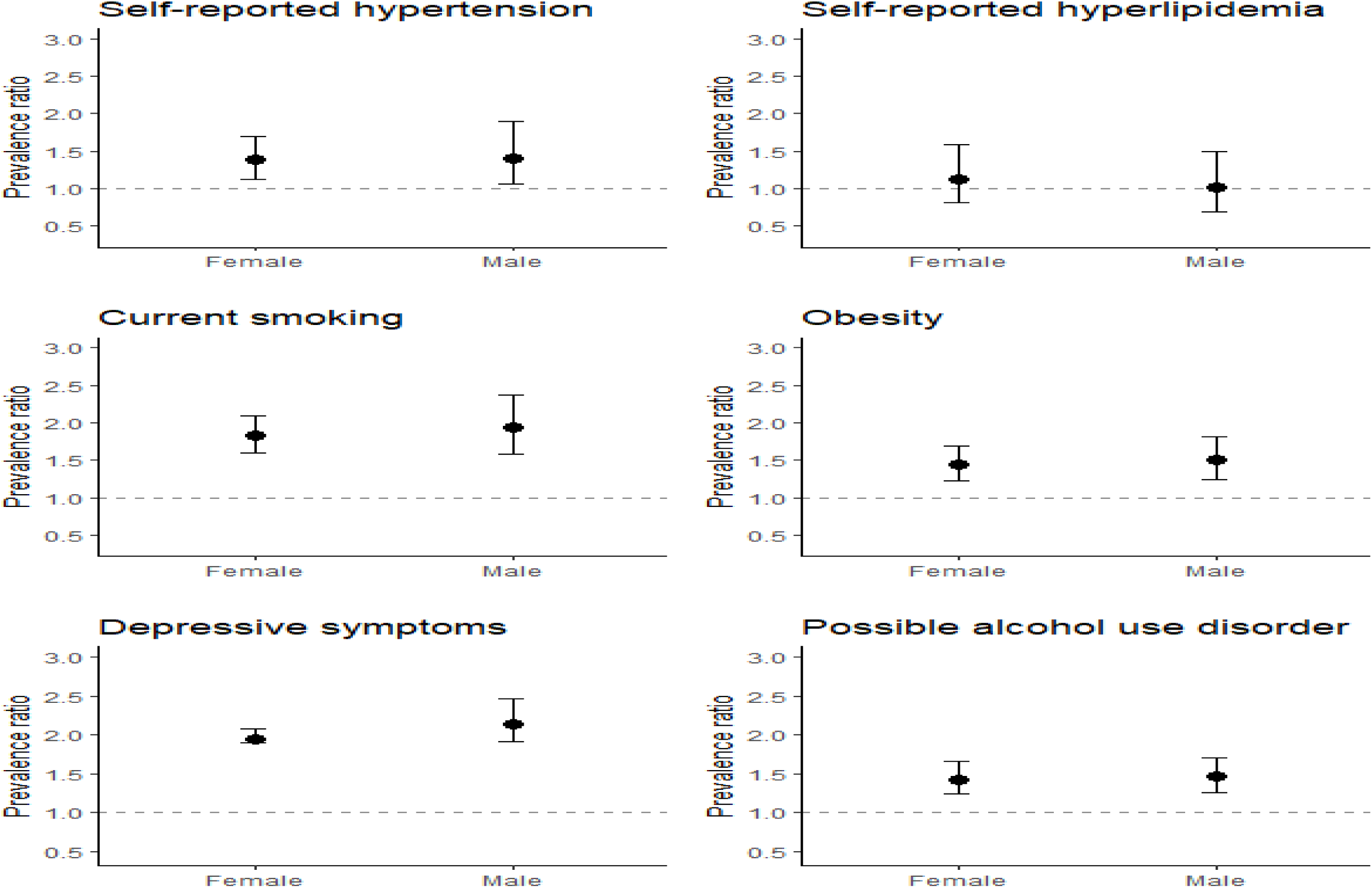
Prevalence ratios of cardiovascular risk factors and mental health indicators comparing those exposed to violence to those not exposed by sex.

The association between lifetime violence exposure and clinical cardiometabolic markers and hs-CRP are presented in Figure 2 and Supplementary Table 4. The absolute mean values were all at the lower end of the normal range, with all mean values rising with age. No statistically significant differences were observed in systolic or diastolic blood pressure by exposure to violence. BMI was higher in individuals exposed to violence (B 0.51; CI: 0.39, 0.63), with more pronounced differences with higher age. The ApoB/A1 ratio was higher in those exposed to violence (B 0.01; CI: 0.01, 0.02) after adjustment, but no difference was seen when examined by sex or age group. Total cholesterol and HbA1c were unchanged by exposure status. Hs-CRP was higher in those exposed to violence (B 0.11; CI: 0.06, 0.16) after multivariable adjustment, with no sex difference.

**Figure 2.**
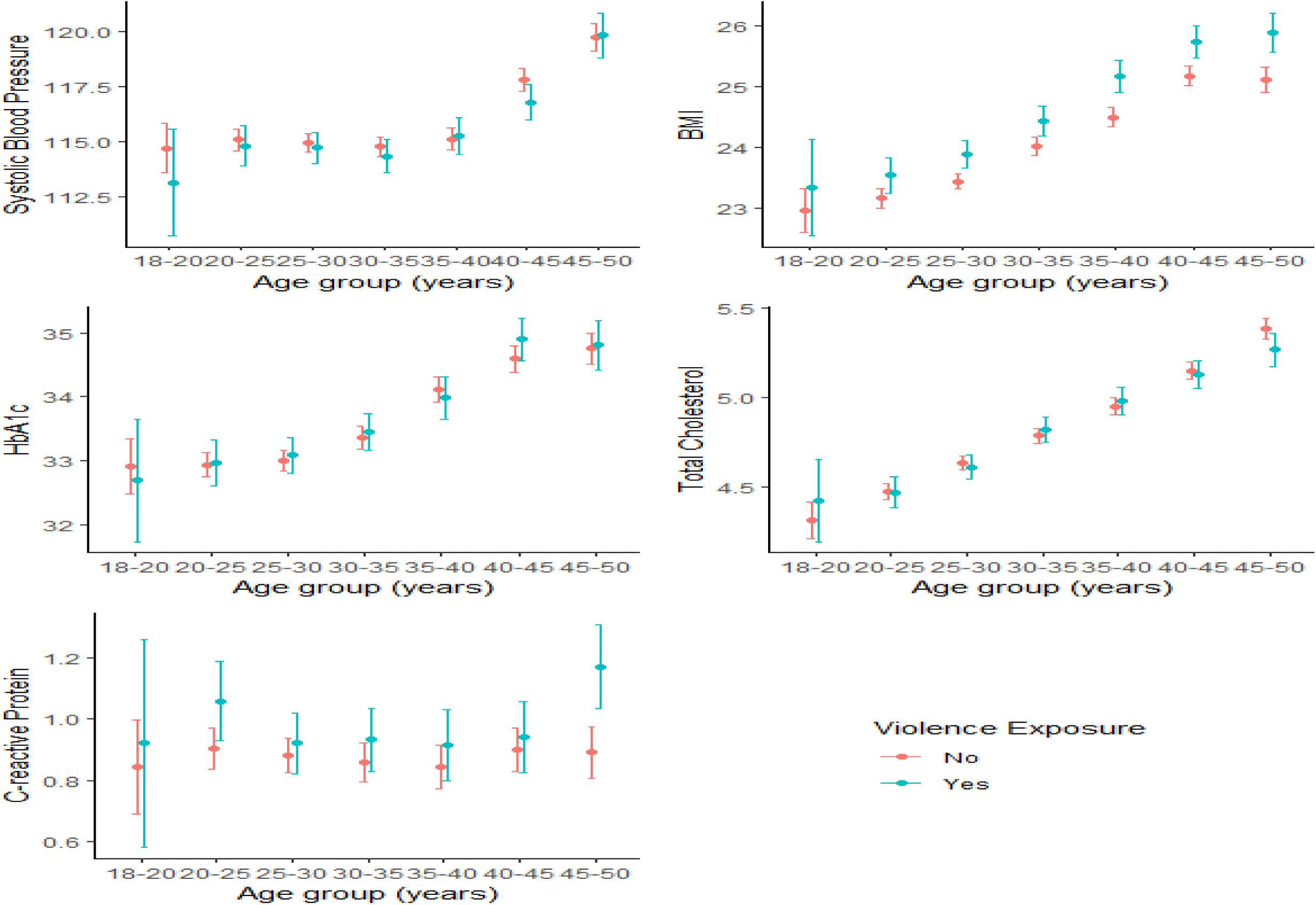
Means with 95% confidence intervals of clinical cardiometabolic markers and C-reactive protein by violence exposure (both sexes) among various age groups.

Tables 2 and 3 as well as Supplementary Tables 5 and 6 show the same outcomes but with age at first exposure, using individuals unexposed to violence as the reference. 3,818 (16.4%) individuals were exposed to violence in this analysis (Supplementary Table 1). Results were similar to the total sample analyses. Additionally, smoking (p = 0.003), obesity (p = 0.01), depressive symptoms (p = 0.02) and hs-CRP (p < 0.001) had stronger associations with violence when first exposed in childhood vs adulthood. These results remained unchanged when stratified by sex (Supplementary Tables 7 and 8).

**Table 2.**
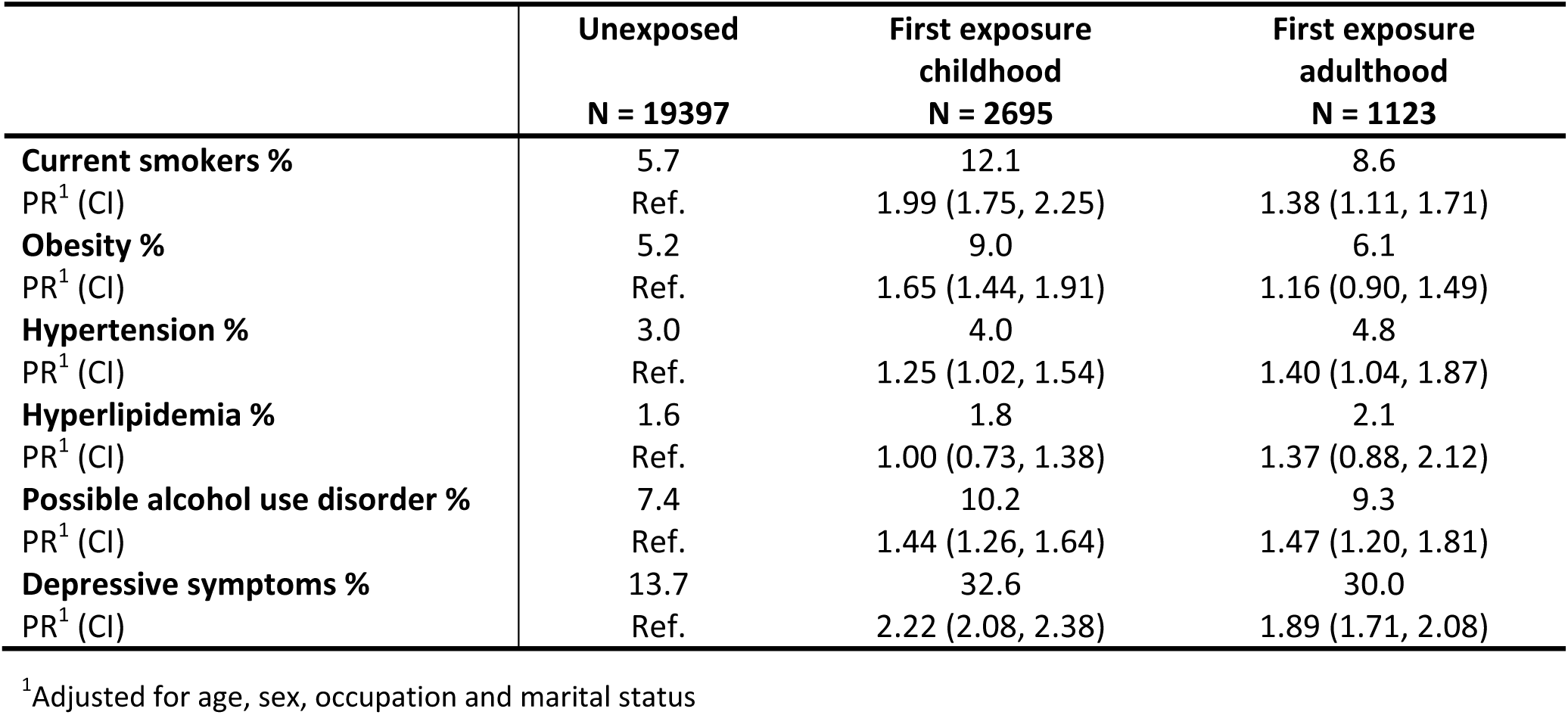
Prevalence ratios of self-reported cardiometabolic diagnoses and mental health indicators by age at first exposure to violence.

**Table 3.** Linear regression of clinical cardiometabolic markers and C-reactive protein by age at first exposure to violence.

| | Unexposed<br>Mean values ± SD<sup>1</sup><br>N = 19397 | First exposure<br>childhood<br>Multivariable difference<sup>2</sup><br>N = 2695 | First exposure<br>adulthood<br>Multivariable difference<sup>2</sup><br>N = 1123 |
| --- | --- | --- | --- |
| **Systolic blood pressure (mmHg)** | 115 ± 13 | -0.40 (-0.83, 0.02) | -0.40 (-1.03, 0.24) |
| **BMI** | 23.9 ± 3.5 | 0.62 (0.48, 0.75) | 0.35 (0.15, 0.56) |
| **Total cholesterol (mmol/L)** | 4.7 ± 0.9 | 0.01 (-0.04, 0.06) | -0.05 (-0.11, 0.02) |
| **HbA1c (mmol/mol)** | 33.5 ± 3.5 | -0.08 (-0.28, 0.13) | 0.24 (-0.04, 0.51) |
| **hs-CRP (mg/L)** | 0.9 ± 1.3 (ref.) | 0.10 (0.04, 0.16) | -0.05 (-0.14, 0.03) |

<sup>1</sup>Standard deviation
<sup>2</sup>Adjusted for age, sex, occupation and marital status

Finally, the analyses by type of violence exposure (Supplementary Tables 9 and 10) showed that exposure to both physical and sexual violence had a stronger association with smoking and depressive symptoms, compared with exposure to only physical or only sexual violence (both p < 0.001). No differences by type of violence exposure were seen with regards to hs-CRP nor BMI.

## Discussion

In a sample of relatively healthy Swedes, aged 18-50, we found that both males and females exposed to violence during their life course had an increased prevalence of smoking, probable alcohol use disorder and depressive symptoms as well as higher BMI and hs-CRP levels, a marker of inflammation, compared to their counterparts without such exposure. These increases were generally more pronounced among those exposed in childhood, as well as among individuals exposed to both physical and sexual violence.

The findings of our study add to a growing body of evidence linking violence to cardiovascular disease[30, 31], and its early markers[31]. BMI was increased in all exposure categories, especially among those exposed in childhood and the differences between those exposed and non-exposed to violence increased with age. This was observed despite the fact that the mean BMI of the study population was in the healthy weight range. Our results on BMI are similar to a longitudinal British study for both genders on childhood maltreatment[13]. We also found a higher prevalence of smoking and possible alcohol use disorder among those exposed to violence, which is in tune with previous studies reporting higher risks of smoking[8] and harmful alcohol use[8] among victims of violence. These negative health behaviors are also associated with depression[32, 33], symptoms of which were also increased in our study among those exposed to violence.

The pathophysiology from violence to cardiovascular disease is still being elucidated. One probable pathway is through inflammation, with increased levels of CRP representing one possible underlying mechanism[34]. Although the protein itself is not causally linked to coronary heart disease[35], it is a predictor of it[36]. A 2016 review of 18 studies on childhood trauma and hs-CRP with a total of 16,870 participants (both genders) found an overall positive association with hs-CRP, though not when physical and sexual abuse were examined specifically[37]. However, there was great heterogeneity and relatively small sample sizes in the studies included in the review. The study population of the present study is indeed larger than all the studies included in the review[37]. A more recent review on childhood maltreatment and inflammation found a positive association for CRP, but only in prospective studies[38]. A small study on intimate partner violence among women found higher CRP among those exposed to violence[20]. Another study found mixed results for sexual abuse in childhood/adolescence using the Nurses’ Health Study II[19]. As CRP and BMI are highly correlated[39], it is not surprising that both measures showed an association with exposure to violence in this study.

Interestingly, despite different patterns of violence exposure by sex in our sample, the associations between violence exposure and cardiometabolic risk factors were largely the same. We expected more differences to arise as biological sex, as well as gender, are a strong determinant of cardiovascular disease[40]. Additionally, females have a different make up of modifiable risk factors[2], and for them, smoking and diabetes may be more hazardous compared with males[41]. Data on sex differences after violence exposure come mainly from childhood abuse studies though the American Heart Association has encouraged even more research into sex differences in childhood adversity and cardiometabolic outcomes[42].

A recent study comparing the LifeGene cohort with data collected by the Swedish Public Health Agency found the LifeGene cohort to be younger, with more non-smokers, and to have a higher educational level[43]. Indeed, our study population proved to be physically healthy, with optimal blood pressure, and low average BMI[44] which is reflected in the overall low prevalence of cardiometabolic diagnoses. Hypertension was the only self-reported diagnosis with a raised prevalence among those exposed to violence, although differences in blood pressure measurements were not statistically significant. This adds to the literature demonstrating increased self-reported hypertension[14, 15], but not measured blood pressure[7], in relation to exposure to violence. One possibility is that this discrepancy is due to the efficacy of hypertensive medications; though the European Society of Cardiology generally aims for blood pressure <130/80[45] which is much higher than the mean blood pressure in the exposed group (114.8 ± 12.4mmHg) of the present study.

Neither cardiometabolic diagnoses, nor their measured counterparts, HbA1c, total cholesterol, and ApoB/A1 ratio, differed by exposure to violence overall. Although null results were observed, a positive association later in life cannot be ruled out[13, 16]. Another possible explanation is the lower prevalence of violence than expected in the present study. A recent Swedish survey of 10,337 persons presented considerably higher prevalences of exposure to violence[46].

To our knowledge, this is the first paper examining lifetime exposure to violence and cardiometabolic outcomes using both sexes. Most studies have focused on women, but there are studies examining childhood exposure and intimate partner violence separately in both sexes. A study of 40-69 year olds from the U.K. Biobank found that depressive/anxiety symptoms, CRP, BMI and smoking mediated the relationship between both physical and sexual abuse in childhood and cardiovascular disease[47]. This study did not find a sex difference in the association between abuse and each chosen mediator but found that the contribution of each mediator towards cardiovascular disease differed by sex. Two large studies using data from the US-based Add Health study, have examined cardiovascular risk profiles in young adults who have experienced intimate partner violence[21, 48]. Both found a small, but increased 30-year risk. A mediation analysis of the same dataset found depressive symptoms, but not alcohol dependence, to mediate the association between exposure to intimate partner violence in the last year and increased cardiovascular risk[49], two variables that were associated with violence exposure in our sample.

### Strengths and limitations

A clear strength of this study is the large sample size, 23,215 males and females; with 80% donating a blood sample. Another strength is the broad data collection, with both self-reported diagnoses and a pool of potential covariates collected in the online questionnaire as well as a clinical visit with objective cardiometabolic markers, that do not suffer the possible bias present in self-reported outcomes. This in turn yields a more selected, healthy population as this commitment is perhaps more feasible for those less affected by violence. Another limitation is the retrospective reporting of violence exposure, though most studies rely on this method, and has a moderate concordance with prospective reporting[50]. To combat this, we used a validated instrument for violence exposure[51].

### Conclusion

The finding of increased BMI and hs-CRP levels in healthy individuals exposed to violence in a Swedish sample of 23,215 indicates very early signals of adverse cardiometabolic health following exposure to violence, with minor sex differences. This adds to the literature that exposure to violence may be associated with cardiometabolic risk factors, including in males, an underrecognized demographic, and that further research in the possible pathways is needed.

## Data Availability

Data availability is subject to ethical approval by the Swedish Ethical Review Authority. Applicants must be associated with a university or research institute based in Sweden, see further at https://lifegene.se/

## Supplementary Tables

**Supplementary Table 1.** List of questions in the modified Life Stressor Checklist-Revised on violence and no. of individuals responding yes to each question.

|  | All<br>Yes (%) | Females<br>Yes (%) | Males<br>Yes (%) |
| --- | --- | --- | --- |
| **Experienced physical violence** | 2916 (12.6) | 1630 (11.6) | 1286 (14.0) |
| Been a victim of robbery or mugging | 1209 (5.2) | 489 (3.5) | 720 (7.8) |
| Before age 18, have you been victimized or abused (not sexually) by someone you knew? | 1382 (6.0) | 757 (5.4) | 625 (6.8) |
| As an adult, have you been victimized or abused (not sexually) by someone you knew? | 802 (3.5) | 620 (4.4) | 182 (2.0) |
| **Experienced sexual violence** | 2449 (10.5) | 2218 (15.8) | 231 (2.5) |
| Before age 18, have you been subjected to sexual harassment or forced to perform sexual acts against your will? | 1541 (6.6) | 1360 (9.7) | 181 (2.0) |
| As an adult, have you been subjected to sexual harassment or forced to perform sexual acts against your will? | 928 (4.0) | 888 (6.3) | 40 (0.4) |
| Before age 18, have you been forced to have sex? | 381 (1.6) | 345 (2.5) | 36 (0.4) |
| As an adult, have you been forced to have sex? | 431 (1.9) | 411 (2.9) | 20 (0.2) |

**Supplementary Table 2.** Background characteristics of males and females in the Swedish LifeGene Cohort by availability of blood test.

|  | **All** |  | **Females** |  | **Males** |  |
| --- | --- | --- | --- | --- | --- | --- |
|  | **No blood test**<br>N = 5052 | **Blood test**<br>N = 18163 | **No blood test**<br>N = 3083 | **Blood test**<br>N = 10942 | **No blood test**<br>N = 1969 | **Blood test**<br>N = 7221 |
| **Age, mean (SD)<sup>1</sup>** | 33.1 (7.7) | 33.1 (8.2) | 32.8 (7.8) | 32.8 (8.4) | 33.6 (7.5) | 33.6 (7.9) |
| Information missing | 0 (0.0) | 0 (0.0) | 0 (0.0) | 0 (0.0) | 0 (0.0) | 0 (0.0) |
| **Education level (%)** |  |  |  |  |  |  |
| Primary | 77 (1.5) | 212 (1.2) | 43 (1.4) | 102 (0.9) | 34 (1.7) | 110 (1.5) |
| Secondary | 1701 (33.7) | 4871 (26.8) | 976 (31.7) | 2809 (25.7) | 725 (36.8) | 2062 (28.6) |
| University | 3173 (62.8) | 12903 (71.0) | 2013 (65.3) | 7929 (72.5) | 1160 (58.9) | 4974 (68.9) |
| Information missing | 101 (2.0) | 177 (1.0) | 51 (1.7) | 102 (0.9) | 50 (2.5) | 75 (1.0) |
| **Occupation (%)** |  |  |  |  |  |  |
| Employed | 3501 (69.3) | 12634 (69.6) | 2080 (67.5) | 7364 (67.3) | 1421 (72.2) | 5270 (73.0) |
| Student | 726 (14.4) | 2783 (15.3) | 489 (15.9) | 1812 (16.6) | 237 (12.0) | 971 (13.4) |
| Other<sup>2</sup> | 295 (5.8) | 788 (4.3) | 191 (6.2) | 524 (4.8) | 104 (5.3) | 264 (3.7) |
| Information missing | 535 (10.3) | 1958 (10.8) | 323 (10.5) | 1242 (11.4) | 207 (10.5) | 716 (9.9) |
| **Civil status (%)** |  |  |  |  |  |  |
| Marriage/Cohabitation | 3150 (62.4) | 10772 (59.3) | 1906 (61.8) | 6478 (59.2) | 1244 (63.2) | 4294 (59.5) |
| Relationship | 469 (9.3) | 1817 (10.0) | 313 (10.2) | 1128 (10.3) | 156 (7.9) | 689 (9.5) |
| Single/Widow | 1303 (25.8) | 5272 (29.0) | 796 (25.8) | 3156 (28.8) | 507 (25.7) | 2116 (29.3) |
| Information missing | 130 (2.6) | 302 (1.7) | 68 (2.2) | 180 (1.6) | 62 (3.1) | 122 (1.7) |

<sup>1</sup> Standard deviation
<sup>2</sup> Unemployed, disability, retired, homemaker or sick leave

**Supplementary Table 3.** Prevalence ratios of self-reported cardiometabolic diagnoses and mental health indicators as well as obesity by sex and violence exposure.

|  | All |  | Females |  | Males |  |
| --- | --- | --- | --- | --- | --- | --- |
|  | Unexposed<br>N = 18527 | Violence<br>N = 4688 | Unexposed<br>N = 10757 | Violence<br>N = 3268 | Unexposed<br>N = 7770 | Violence<br>N = 1420 |
| **Current smokers %** | 5.5 | 10.8 | 5.9 | 11.0 | 4.8 | 10.4 |
| PR<sup>1</sup> (CI) | Ref. | 1.94 (1.75, 2.15) | Ref. | 1.88 (1.66, 2.12) | Ref. | 2.10 (1.75, 2.52) |
| PR<sup>2</sup> (CI) | Ref. | 1.86 (1.66, 2.07) | Ref. | 1.83 (1.60, 2.09) | Ref. | 1.94 (1.59, 2.37) |
| **Obesity %** | 5.2 | 7.9 | 4.6 | 7.0 | 6.0 | 10.1 |
| PR<sup>1</sup> (CI) | Ref. | 1.50 (1.33, 1.68) | Ref. | 1.42 (1.22, 1.65) | Ref. | 1.61 (1.35, 1.92) |
| PR<sup>2</sup> (CI) | Ref. | 1.46 (1.30, 1.65) | Ref. | 1.44 (1.23, 1.69) | Ref. | 1.50 (1.24, 1.82) |
| **Hypertension %** | 2.9 | 4.4 | 3.0 | 4.5 | 2.8 | 4.3 |
| PR<sup>1</sup> (CI) | Ref. | 1.41 (1.20, 1.65) | Ref. | 1.39 (1.15, 1.68) | Ref. | 1.45 (1.10, 1.92) |
| PR<sup>2</sup> (CI) | Ref. | 1.39 (1.18, 1.64) | Ref. | 1.38 (1.13, 1.69) | Ref. | 1.41 (1.06, 1.89) |
| **Diabetes type 2 %** | 0.1 | 0.2 | 0.1 | 0.2 | 0.1 | 0.2 |
| PR<sup>1</sup> (CI) | Ref. | 1.36 (0.62, 2.97) | Ref. | 1.22 (0.47, 3.21) | Ref. | 1.69 (0.46, 6.14) |
| PR<sup>2</sup> (CI) | Ref. | 1.19 (0.53, 2.69) | Ref. | 1.10 (0.40, 3.03) | Ref. | 1.38 (0.37, 5.15) |
| **Hyperlipidemia %** | 1.6 | 1.9 | 0.8 | 1.6 | 2.2 | 2.5 |
| PR<sup>1</sup> (CI) | Ref. | 1.14 (0.90, 1.44) | Ref. | 1.22 (0.89, 1.67) | Ref. | 1.06 (0.74, 1.50) |
| PR<sup>2</sup> (CI) | Ref. | 1.07 (0.84, 1.37) | Ref. | 1.13 (0.81, 1.58) | Ref. | 1.02 (0.69, 1.49) |
| **Possible alcohol use disorder %** | 7.1 | 10.3 | 5.5 | 8.5 | 9.4 | 14.4 |
| PR<sup>1</sup> (CI) | Ref. | 1.57 (1.42, 1.73) | Ref. | 1.58 (1.38, 1.81) | Ref. | 1.56 (1.35, 1.80) |
| PR<sup>2</sup> (CI) | Ref. | 1.45 (1.30, 1.61) | Ref. | 1.43 (1.24, 1.66) | Ref. | 1.47 (1.26, 1.71) |
| **Depressive symptoms %** | 13.6 | 28.8 | 16.9 | 33.0 | 9.1 | 19.2 |
| PR<sup>1</sup> (CI) | Ref. | 2.05 (1.94, 2.17) | Ref. | 2.02 (1.90, 2.15) | Ref. | 2.17 (1.91, 2.46) |
| PR<sup>2</sup> (CI) | Ref. | 1.99 (1.87, 2.11) | Ref. | 1.95 (1.82, 2.08) | Ref. | 2.13 (1.87, 2.43) |

<sup>1</sup>Age adjusted, with all column also adjusted for sex
<sup>2</sup>Adjusted for age, occupation and marital status, with all column also adjusted for sex

**Supplementary Table 4.**
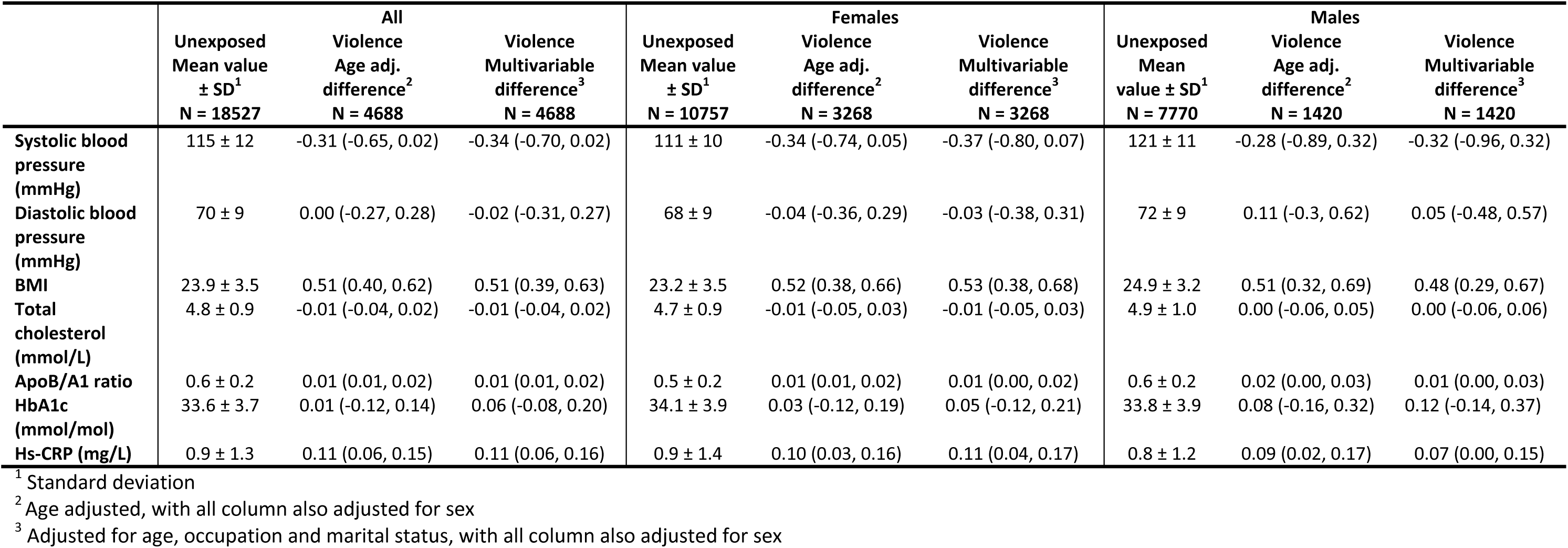
Linear regression of clinical cardiometabolic markers and C-reactive protein by sex and violence exposure.

|  | All |  |  | Females |  |  | Males |  |  |
| --- | --- | --- | --- | --- | --- | --- | --- | --- | --- |
|  | Unexposed<br>Mean value<br>± SD<sup>1</sup><br>N = 18527 | Violence<br>Age adj.<br>difference<sup>2</sup><br>N = 4688 | Violence<br>Multivariable<br>difference<sup>3</sup><br>N = 4688 | Unexposed<br>Mean value<br>± SD<sup>1</sup><br>N = 10757 | Violence<br>Age adj.<br>difference<sup>2</sup><br>N = 3268 | Violence<br>Multivariable<br>difference<sup>3</sup><br>N = 3268 | Unexposed<br>Mean<br>value ± SD<sup>1</sup><br>N = 7770 | Violence<br>Age adj.<br>difference<sup>2</sup><br>N = 1420 | Violence<br>Multivariable<br>difference<sup>3</sup><br>N = 1420 |
| **Systolic blood pressure (mmHg)** | 115 ± 12 | -0.31 (-0.65, 0.02) | -0.34 (-0.70, 0.02) | 111 ± 10 | -0.34 (-0.74, 0.05) | -0.37 (-0.80, 0.07) | 121 ± 11 | -0.28 (-0.89, 0.32) | -0.32 (-0.96, 0.32) |
| **Diastolic blood pressure (mmHg)** | 70 ± 9 | 0.00 (-0.27, 0.28) | -0.02 (-0.31, 0.27) | 68 ± 9 | -0.04 (-0.36, 0.29) | -0.03 (-0.38, 0.31) | 72 ± 9 | 0.11 (-0.3, 0.62) | 0.05 (-0.48, 0.57) |
| **BMI** | 23.9 ± 3.5 | 0.51 (0.40, 0.62) | 0.51 (0.39, 0.63) | 23.2 ± 3.5 | 0.52 (0.38, 0.66) | 0.53 (0.38, 0.68) | 24.9 ± 3.2 | 0.51 (0.32, 0.69) | 0.48 (0.29, 0.67) |
| **Total cholesterol (mmol/L)** | 4.8 ± 0.9 | -0.01 (-0.04, 0.02) | -0.01 (-0.04, 0.02) | 4.7 ± 0.9 | -0.01 (-0.05, 0.03) | -0.01 (-0.05, 0.03) | 4.9 ± 1.0 | 0.00 (-0.06, 0.05) | 0.00 (-0.06, 0.06) |
| **ApoB/A1 ratio** | 0.6 ± 0.2 | 0.01 (0.01, 0.02) | 0.01 (0.01, 0.02) | 0.5 ± 0.2 | 0.01 (0.01, 0.02) | 0.01 (0.00, 0.02) | 0.6 ± 0.2 | 0.02 (0.00, 0.03) | 0.01 (0.00, 0.03) |
| **HbA1c (mmol/mol)** | 33.6 ± 3.7 | 0.01 (-0.12, 0.14) | 0.06 (-0.08, 0.20) | 34.1 ± 3.9 | 0.03 (-0.12, 0.19) | 0.05 (-0.12, 0.21) | 33.8 ± 3.9 | 0.08 (-0.16, 0.32) | 0.12 (-0.14, 0.37) |
| **Hs-CRP (mg/L)** | 0.9 ± 1.3 | 0.11 (0.06, 0.15) | 0.11 (0.06, 0.16) | 0.9 ± 1.4 | 0.10 (0.03, 0.16) | 0.11 (0.04, 0.17) | 0.8 ± 1.2 | 0.09 (0.02, 0.17) | 0.07 (0.00, 0.15) |

<sup>1</sup> Standard deviation
<sup>2</sup> Age adjusted, with all column also adjusted for sex
<sup>3</sup> Adjusted for age, occupation and marital status, with all column also adjusted for sex

**Supplementary Table 5.**
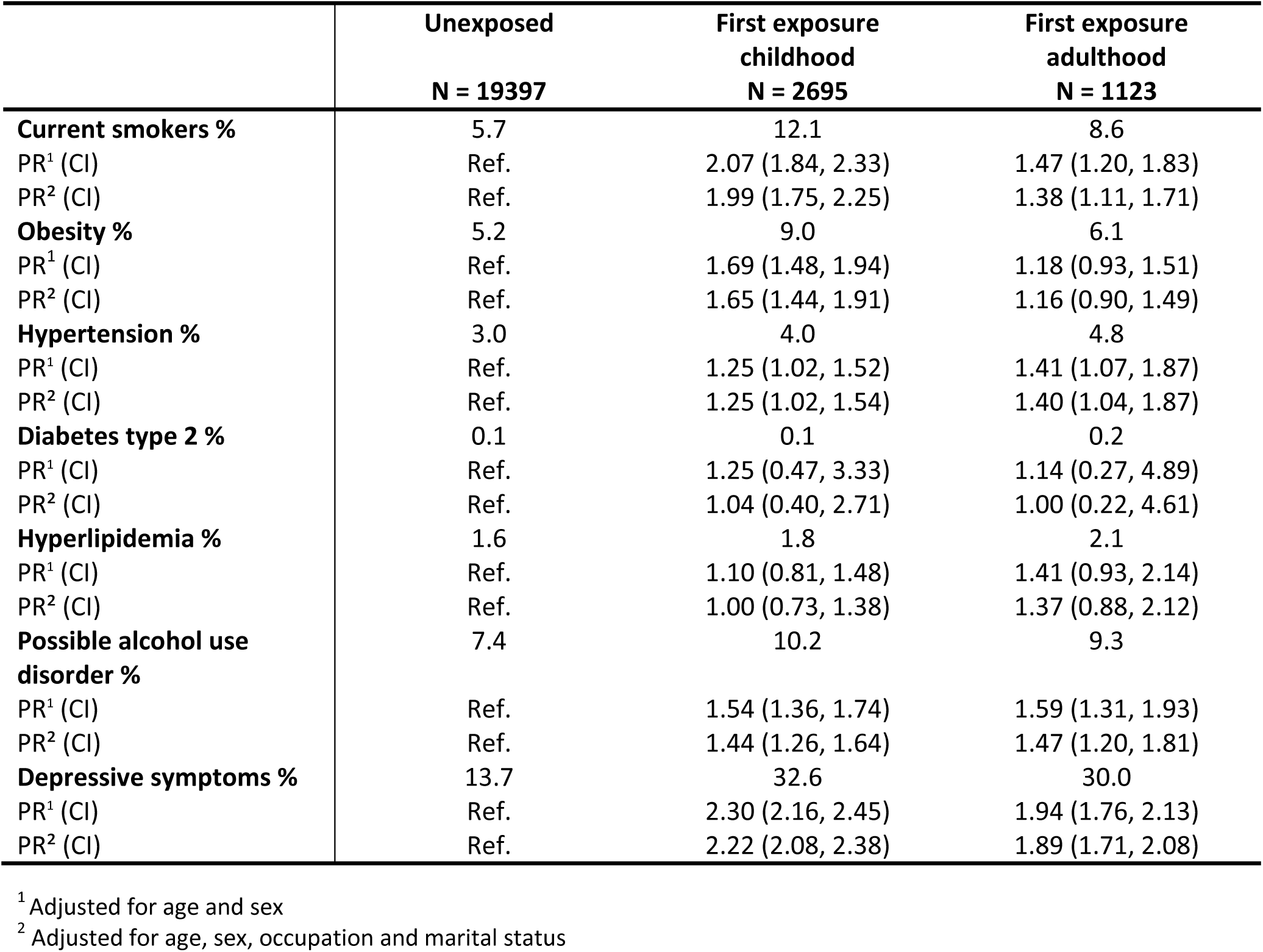
Prevalence ratios of self-reported cardiometabolic diagnoses and mental health indicators by age at first exposure to violence.

**Supplementary Table 6.** Linear regression of clinical cardiometabolic markers and C-reactive protein by age at first exposure to violenc.

|  | **Unexposed** | **First exposure**<br>**childhood** | **First exposure**<br>**adulthood** | **First exposure**<br>**childhood** | **First exposure**<br>**adulthood** |
| --- | --- | --- | --- | --- | --- |
|  | **Mean values ± SD<sup>1</sup>** | **Age adj. difference<sup>2</sup>** | **Age adj. difference<sup>2</sup>** | **Multivariable difference<sup>3</sup>** | **Multivariable difference<sup>3</sup>** |
|  | **N = 19397** | **N = 2695** | **N = 1123** | **N = 2695** | **N = 1123** |
| **Systolic blood pressure (mmHg)** | 115 ± 13 | -0.40 (-0.83, 0.02) | -0.40 (-1.03, 0.24) | -0.40 (-0.85, 0.06) | -0.44 (-1.12, 0.24) |
| **Diastolic blood pressure (mmHg)** | 70 ± 9 | 0.06 (-0.29, 0.40) | 0.02 (-0.50, 0.54) | 0.04 (-0.32, 0.41) | -0.03 (-0.58, 0.52) |
| **BMI** | 23.9 ± 3.5 | 0.62 (0.48, 0.75) | 0.35 (0.15, 0.56) | 0.61 (0.46, 0.76) | 0.40 (0.18, 0.62) |
| **Total cholesterol (mmol/L)** | 4.8 ± 0.9 | 0.01 (-0.03, 0.05) | -0.01 (-0.12, -0.01) | 0.01 (-0.03, 0.05) | -0.01 (-0.13, 0.00) |
| **ApoB/A1 ratio** | 0.6 ± 0.2 | 0.02 (0.01, 0.03) | 0.00 (-0.01, 0.01) | 0.02 (0.01, 0.02) | 0.00 (-0.01, 0.02) |
| **HbA1c (mmol/mol)** | 33.6 ± 3.7 | -0.02 (-0.18, 0.15) | 0.02 (-0.22, 0.27) | 0.02 (-0.16, 0.20) | 0.19 (-0.07, 0.46) |
| **hs-CRP (mg/L)** | 0.9 ± 1.3 | 0.10 (0.04, 0.16) | -0.05 (-0.14, 0.03) | 0.16 (0.10, 0.22) | -0.04 (-0.13, 0.06) |

<sup>1</sup> Standard deviation
<sup>2</sup> Adjusted for age and sex
<sup>3</sup> Adjusted for age, sex, occupation and marital status

**Supplementary Table 7.** Prevalence ratios of self-reported cardiometabolic diagnoses and mental health indicators by sex and age at first exposure to violence.

|  | Females |  |  | Males |  |  |
| --- | --- | --- | --- | --- | --- | --- |
|  | Unexposed<br>N = 11079 | First exposure<br>childhood<br>N = 1939 | First exposure<br>adulthood<br>N = 1007 | Unexposed<br>N = 8318 | First exposure<br>childhood<br>N = 756 | First exposure<br>adulthood<br>N = 116 |
| **Current smokers %** | 6.0 | 12.6 | 8.2 | 5.1 | 10.7 | 12.1 |
| PR<sup>1</sup> (CI) | Ref. | 2.06 (1.80, 2.37) | 1.41 (1.13, 1.75) | Ref. | 2.07 (1.65, 2.59) | 2.14 (1.30, 3.52) |
| PR<sup>2</sup> (CI) | Ref. | 2.04 (1.77, 2.37) | 1.32 (1.05, 1.67) | Ref. | 1.84 (1.43, 2.37) | 1.97 (1.17, 3.33) |
| **Obesity %** | 4.6 | 8.0 | 5.5 | 6.1 | 11.6 | 12.1 |
| PR<sup>1</sup> (CI) | Ref. | 1.62 (1.37, 1.93) | 1.06 (0.81, 1.39) | Ref. | 1.80 (1.46, 2.22) | 1.87 (1.13, 3.10) |
| PR<sup>2</sup> (CI) | Ref. | 1.66 (1.39, 1.99) | 1.08 (0.81, 1.43) | Ref. | 1.66 (1.32, 2.08) | 1.65 (0.95, 2.87) |
| **Hypertension %** | 3.1 | 4.1 | 4.7 | 2.9 | 4.0 | 6.0 |
| PR<sup>1</sup> (CI) | Ref. | 1.24 (0.98, 1.58) | 1.36 (1.01, 1.84) | Ref. | 1.26 (0.87, 1.82) | 1.91 (0.92, 3.97) |
| PR<sup>2</sup> (CI) | Ref. | 1.26 (0.98, 1.61) | 1.37 (1.00, 1.87) | Ref. | 1.23 (0.84, 1.80) | 1.79 (0.81, 3.96) |
| **Diabetes type 2 %** | 0.1 | 0.2 | 0.2 | 0.1 | 0.3 | 0.0 |
| PR<sup>1</sup> (CI) | Ref. | 0.99 (0.28, 3.50) | 1.22 (0.27, 5.37) | Ref. | 1.93 (0.43, 8.67) | 0.00 (0.00, 0.00) |
| PR<sup>2</sup> (CI) | Ref. | 0.86 (0.26, 2.88) | 1.09 (0.22, 5.36) | Ref. | 1.44 (0.32, 6.50) | 0.00 (0.00, 0.00) |
| **Hyperlipidemia %** | 1.2 | 1.5 | 2.0 | 1.6 | 1.8 | 3.4 |
| PR<sup>1</sup> (CI) | Ref. | 1.18 (0.80, 1.75) | 1.44 (0.91, 2.31) | Ref. | 1.00 (0.63, 1.59) | 1.39 (0.53, 3.65) |
| PR<sup>2</sup> (CI) | Ref. | 1.05 (0.69, 1.59) | 1.40 (0.86, 2.27) | Ref. | 0.97 (0.59, 1.61) | 1.22 (0.40, 3.76) |
| **Possible alcohol use disorder %** | 5.6 | 8.7 | 8.4 | 9.7 | 14.2 | 16.5 |
| PR<sup>1</sup> (CI) | Ref. | 1.58 (1.34, 1.86) | 1.56 (1.26, 1.94) | Ref. | 1.49 (1.24, 1.80) | 1.76 (1.16, 2.66) |
| PR<sup>2</sup> (CI) | Ref. | 1.46 (1.23, 1.75) | 1.42 (1.13, 1.80) | Ref. | 1.41 (1.16, 1.72) | 1.71 (1.10, 2.63) |
| **Depressive symptoms %** | 17.0 | 36.0 | 31.4 | 9.3 | 23.8 | 18.1 |
| PR<sup>1</sup> (CI) | Ref. | 2.21 (2.06, 2.37) | 1.91 (1.73, 2.10) | Ref. | 2.66 (2.31, 3.06) | 2.11 (1.44, 3.09) |
| PR<sup>2</sup> (CI) | Ref. | 2.15 (1.99, 2.31) | 1.86 (1.68, 2.06) | Ref. | 2.51 (2.17, 2.91) | 2.17 (1.49, 3.15) |

<sup>1</sup> Adjusted for age
<sup>2</sup> Adjusted for age, occupation and marital status

**Supplementary Table 8.** Linear regression of clinical cardiometabolic markers and C-reactive protein by age at first exposure to violence.

|  | Females |  |  |  |  | Males |  |  |  |  |
| --- | --- | --- | --- | --- | --- | --- | --- | --- | --- | --- |
| | Unexposed<br>Mean<br>value ± SD<sup>1</sup><br>N = 11079 | First exposure<br>childhood<br>Age adj.<br>difference<sup>2</sup><br>N = 1939 | First exposure<br>adulthood<br>Age adj.<br>difference<sup>2</sup><br>N = 1007 | First exposure<br>childhood<br>Multivariable<br>difference<sup>3</sup><br>N = 1939 | First exposure<br>adulthood<br>Multivariable<br>difference<sup>3</sup><br>N = 1007 | Unexposed<br>Mean<br>value ± SD<sup>1</sup><br>N = 8318 | First exposure<br>childhood<br>Age adj.<br>difference<sup>2</sup><br>N = 756 | First exposure<br>adulthood<br>Age adj.<br>difference<sup>2</sup><br>N = 116 | First exposure<br>childhood<br>Multivariable<br>difference<sup>3</sup><br>N = 756 | First exposure<br>adulthood<br>Multivariable<br>difference<sup>3</sup><br>N = 116 |
| **Systolic blood pressure (mmHg)** | 111 ± 11 | -0.23 (-0.73, 0.27) | -0.45 (-1.12, 0.22) | -0.31 (-0.85, 0.22) | -0.43 (-1.15, 0.28) | 120 ± 11 | -0.80 (-1.61, 0.00) | -0.95 (-2.92, 1.02) | -0.67 (-1.51, 0.17) | -1.47 (-3.56, 0.63) |
| **Diastolic blood pressure (mmHg)** | 68 ± 9 | -0.04 (-0.44, 0.36) | 0.08 (-0.46, 0.61) | -0.05 (-0.48, 0.38) | 0.04 (-0.53, 0.61) | 72 ± 9 | 0.31 (-0.36, 0.97) | -0.26 (-1.89, 1.38) | 0.26 (-0.42, 0.96) | -0.42 (-2.15, 1.30) |
| **BMI** | 23.2 ± 3.5 | 0.65 (0.48, 0.82) | 0.27 (0.04, 0.49) | 0.65 (0.47, 0.84) | 0.32 (0.08, 0.57) | 24.9 ± 3.2 | 0.54 (0.30, 0.77) | 1.11 (0.52, 1.70) | 0.52 (0.27, 0.77) | 1.03 (0.41, 1.65) |
| **Total cholesterol (mmol/L)** | 4.7 ± 0.9 | 0.01 (-0.03, 0.06) | -0.04 (-0.10, 0.02) | 0.01 (-0.04, 0.06) | -0.05 (-0.11, 0.02) | 4.9 ± 1.0 | 0.02 (-0.06, 0.10) | -0.13 (-0.32, -0.06) | 0.02 (-0.06, 0.10) | -0.14 (-0.34, 0.06) |
| **ApoB/A1 ratio** | 0.5 ± 0.2 | 0.02 (0.01, 0.03) | 0.00 (-0.01, 0.01) | 0.02 (0.01, 0.03) | 0.00 (-0.01, 0.01) | 0.6 ± 0.2 | 0.02 (0.00, 0.03) | 0.04 (0.00, 0.08) | 0.01 (0.00, 0.03) | 0.03 (-0.01, 0.08) |
| **HbA1c (mmol/mol)** | 33.5 ± 3.5 | -0.05 (-0.24, 0.14) | 0.15 (-0.11, 0.40) | -0.08 (-0.28, 0.13) | 0.24 (-0.04, 0.51) | 33.8 ± 3.9 | 0.22 (-0.09, 0.53) | -0.31 (-1.09, 0.47) | 0.27 (-0.07, 0.60) | -0.31 (-1.14, 0.53) |
| **hs-CRP (mg/L)** | 0.9 ± 1.4 | 0.15 (0.07, 0.22) | -0.03 (-0.14, 0.06) | 0.17 (0.08, 0.25) | -0.04 (-0.14, 0.07) | 0.8 ± 1.2 | 0.17 (0.07, 0.26) | 0.07 (-0.18, 0.31) | 0.15 (0.04, 0.25) | 0.05 (-0.21, 0.32) |

<sup>1</sup> Standard deviation
<sup>2</sup> Adjusted for age
<sup>3</sup> Adjusted for age, occupation and marital status

**Supplementary Table 9.** Prevalence ratios of self-reported cardiometabolic diagnoses and mental health indicators by type of violence exposure.

|  | Unexposed | Physical only | Sexual only | Physical and Sexual |
| --- | --- | --- | --- | --- |
|  | N = 18527 | N = 2239 | N = 1772 | N = 677 |
| **Current smokers %** | 5.5 | 10.6 | 9.4 | 15.5 |
| PR<sup>1</sup> (CI) | Ref. | 1.95 (1.70, 2.23) | 1.63 (1.39, 1.91) | 2.74 (2.27, 3.30) |
| PR<sup>2</sup> (CI) | Ref. | 1.84 (1.59, 2.13) | 1.61 (1.36, 1.90) | 2.57 (2.11, 3.14) |
| **Obesity %** | 5.2 | 8.1 | 7.1 | 9.6 |
| PR<sup>1</sup> (CI) | Ref. | 1.44 (1.24, 1.67) | 1.44 (1.20, 1.73) | 1.85 (1.46, 2.36) |
| PR<sup>2</sup> (CI) | Ref. | 1.43 (1.22, 1.67) | 1.42 (1.17, 1.72) | 1.71 (1.33, 2.21) |
| **Hypertension %** | 2.9 | 5.0 | 3.8 | 4.0 |
| PR<sup>1</sup> (CI) | Ref. | 1.64 (1.34, 2.00) | 1.19 (0.92, 1.53) | 1.21 (0.83, 1.77) |
| PR<sup>2</sup> (CI) | Ref. | 1.60 (1.30, 1.97) | 1.16 (0.89, 1.51) | 1.28 (0.87, 1.87) |
| **Diabetes type 2 %** | 0.1 | 0.2 | 0.2 | 0.3 |
| PR<sup>1</sup> (CI) | Ref. | 1.32 (0.47, 3.75) | 1.17 (0.35, 3.95) | 1.97 (0.46, 8.45) |
| PR<sup>2</sup> (CI) | Ref. | 1.22 (0.42, 3.54) | 1.08 (0.31, 3.63) | 1.36 (0.29, 6.31) |
| **Hyperlipidemia %** | 1.6 | 2.0 | 1.5 | 2.8 |
| PR<sup>1</sup> (CI) | Ref. | 1.06 (0.77, 1.45) | 1.00 (0.67, 1.51) | 1.79 (1.13, 2.84) |
| PR<sup>2</sup> (CI) | Ref. | 1.02 (0.73, 1.42) | 0.96 (0.63, 1.46) | 1.59 (0.95, 2.65) |
| **Possible alcohol use disorder %** | 7.1 | 11.6 | 9.4 | 8.4 |
| PR<sup>1</sup> (CI) | Ref. | 1.55 (1.37, 1.76) | 1.66 (1.41, 1.94) | 1.43 (1.11, 1.85) |
| PR<sup>2</sup> (CI) | Ref. | 1.40 (1.23, 1.61) | 1.60 (1.36, 1.89) | 1.26 (0.95, 1.68) |
| **Depressive symptoms %** | 13.6 | 22.6 | 32.1 | 40.9 |
| PR<sup>1</sup> (CI) | Ref. | 1.78 (1.64, 1.93) | 2.06 (1.91, 2.22) | 2.87 (2.62, 3.15) |
| PR<sup>2</sup> (CI) | Ref. | 1.71 (1.57, 1.87) | 2.03 (1.88, 2.20) | 2.70 (2.44, 2.98) |

<sup>1</sup> Adjusted for age
and sex
<sup>2</sup> Adjusted for age, sex, occupation and marital status

**Supplementary Table 10.** Linear regression of clinical cardiometabolic markers and C-reactive protein by type of violence exposure.

|  | Unexposed<br>Mean values<br>± SD<sup>1</sup><br>N = 18527 | Physical only<br>Age adj. difference<sup>2</sup><br>N = 2239 | Sexual only<br>Age adj.<br>difference<sup>2</sup><br>N = 1772 | Physical and sexual<br>Age adj.<br>difference<sup>2</sup><br>N = 677 | Physical only<br>Multivariable<br>difference<sup>3</sup><br>N = 2239 | Sexual only<br>Multivariable<br>difference<sup>3</sup><br>N = 1772 | Physical and sexual<br>Multivariable<br>difference<sup>3</sup><br>N = 677 |
| --- | --- | --- | --- | --- | --- | --- | --- |
| **Systolic blood pressure (mmHg)** | 115 ± 12 | -0.25 (-0.71, 0.21) | -0.35 (-0.87, 0.17) | -0.44 (-1.25, 0.40) | -0.19 (-0.68, 0.30) | -0.47 (-1.02, 0.08) | -0.54 (-1.40, 0.33) |
| **Diastolic blood pressure (mmHg)** | 70 ± 9 | 0.03 (-0.34, 0.41) | -0.02 (-0.44, 0.41) | -0.06 (-0.72, 0.60) | 0.02 (-0.38, 0.41) | -0.05 (-0.50, 0.40) | -0.08 (-0.78, 0.62) |
| **BMI** | 23.9 ± 3.5 | 0.47 (0.32, 0.62) | 0.48 (0.31, 0.65) | 0.75 (0.48, 1.01) | 0.47 (0.31, 0.63) | 0.50 (0.32, 0.68) | 0.71 (0.43, 1.00) |
| **Total cholesterol (mmol/L)** | 4.8 ± 0.9 | 0.00 (-0.04, 0.04) | -0.03 (-0.08, 0.02) | 0.00 (-0.07, 0.08) | 0.00 (-0.04, 0.05) | -0.03 (-0.08, 0.03) | -0.01 (-0.10, 0.07) |
| **ApoB/A1 ratio** | 0.6 ± 0.2 | 0.01 (0.01, 0.02) | 0.01 (0.00, 0.02) | 0.02 (0.00, 0.03) | 0.01 (0.00, 0.02) | 0.01 (0.00, 0.02) | 0.01 (0.00, 0.03) |
| **HbA1c (mmol/mol)** | 33.6 ± 3.7 | 0.7 (-0.10, 0.25) | -0.09 (-0.28, 0.11) | 0.05 (-0.27, 0.36) | 0.04 (-0.15, 0.23) | 0.07 (-0.15, 0.29) | 0.11 (-0.22, 0.45) |
| **CRP (mg/L)** | 0.9 ± 1.3 | 0.08 (0.01, 0.14) | 0.10 (0.03, 0.17) | 0.21 (0.10, 0.33) | 0.07 (0.00, 0.14) | 0.11 (0.03, 0.19) | 0.21 (0.09, 0.33) |

<sup>1</sup> Standard deviation
<sup>2</sup> Adjusted for age and sex
<sup>3</sup> Adjusted for age, sex, occupation and marital status

